# Elevated blood urea nitrogen to creatinine ratio during hospitalization is associated with 90-day poor outcome in ICH patients

**DOI:** 10.1101/2023.11.18.23298732

**Authors:** Yamin Wei, Wenjing Deng, Yanan Zhao, Huijie Shao

## Abstract

**Background and Purpose:** Dehydration is common in hospitalized patients and associated with poor outcome in ischemic stroke patients. Intracerebral hemorrhage patients use hyperosmolar agents frequently after admission, which may lead to dehydration. Since the blood urea nitrogen to creatinine ratio (BUN/Cr) is an indicator of dehydration, it is unknown whether there is a relationship between BUN/Cr ratio during hospitalization and clinical outcome of intracerebral hemorrhage patients.

**Mehtods:** A total of 211 patients with supratentorial cerebral hemorrhage were included. Clinical data was collected retrospectively. BUN/Cr ratio on day 7 after onset (7dBUN/Cr) was calculated. Poor outcome was defined as 90-day mRS>2. Univariate and multivariate logistic regression analyses were used to determine the relationship between 7dBUN/Cr ratio and 90-day poor outcome. Receiver operating curve was used to determine the best cutoff of 7dBUN/Cr ratio for predicting poor outcome.

**Results:** NIHSS score, hematoma volume and 7dBUN/Cr ratio were independently correlated with 90-day poor outcome. Under receiver operating curve, 7dBUN/Cr ratio exhibited similar prognostic capability, as compared to hematoma volume. The best cutoff for 7dBUN/Cr ratio to predict poor outcome was 22 in the hyperosmolar agents subgroup.

**Conclusions:** Elevated BUN/Cr ratio at day 7 is associated with 90-day poor outcome in ICH patients. Further prospective study will be required to confirm this result and explore the value of BUN/Cr ratio in the application of hyperosmolar agents and hydration therapy.

## 1. INTRODUCTION

Intracerebral hemorrhage (ICH)is a subtype of stroke that accounts for about 10% of stroke in the United States^[1]^, and associated with high mortality and high long-term disability. Despite a variety of treatments, such as blood pressure control, hematoma aspiration, and decompressive craniectomy^[2]^, a significant proportion of patients with ICH have a poor outcome.

Many studies indicate that hydration status significantly affects the prognosis of patients with acute ischemic stroke ^[3][4]^. Dehydration is an independent predictor for outcome after ischemic stroke^[5][6][7][8][8][9][11]^. Admission dehydration is associated with worse discharge outcomes and higher costs in acute ischaemic stroke, but not in hemorrhagic stroke^[12]^. However, dehydration is more common in stroke patients after admission^[6]^, due to the appilication of hyperosmolar agents to control brain edema and elevated intracranial pressure, and progression of neurological deficits after stroke, such as consciousness disturbance and dysphagia. To date, there have been few studies on the correlation between dehydration after admission and functional prognosis of stroke.

The diagnosis of dehydration is subjective and lacks a relatively objective gold standard. A common indicator of dehydration is the blood urea nitrogen to creatinine ratio (BUN/Cr)^[13]^. BUN/Cr ratio has been used in several studies to assess hydration status, yielding consistent results in stroke outcome^[14][15][16][17]^. In the current study, we investigated the association between BUN/Cr ratio on day 7(7dBUN/Cr) after onset and 90-day outcome.

## 2. MATERIAL AND METHODS

### 2.1. Study Population

We retrospectively analyzed the data from September 2021 to January 2023 at the First Affiliated Hospital of Zhengzhou University. We enrolled patients using the following criteria.

Inclusion Criteria: (1) The type of ICH is spontaneous intracerebral hemorrhage; (2) supratentorial cerebral hemorrhage; (3) patients are admitted within 24h from the onset of symptoms; (4) medical records including imaging and laboratory data are complete.

Exclusion Criteria: (1) ICH caused by secondary factors, such as trauma, brain tumor, Cerebral infarction, aneurysm, cerebral arteriovenous malformation, and coagulation disorders; (2) intraventricular hemorrhage and subarachnoid hemorrhage(SAH); (3) surgical interventions, such as hematoma evacuation, stereotactic aspiration, EVD insertion, and lumbar cistern drainage; (4) factors that may influence the BUN/Cr ratio, such as renal insufficiency, congestive heart failure, gastrointestinal hemorrhage, and urinary tract obstruction;(5)Age under 18 years old;(6) history of stroke and residual disability.

### 2.2. Data Collection

We collected information including demographic data(gender, age), adverse life habits(smoking and alcohol consumption), medical history(hypertension, diabetes mellitus, dyslipidemia), medication history, imaging data, clinical score(GCS score, NIHSS score) at admission, and laboratory data which included blood routine, liver and kidney function, blood glucose, electrolytes, lipids within 24h after admission, and kidney function on day 7 after onset. Medication history included anticoagulant/antiplatelet drug which had been used for 3 months before the onset, and hyperosmolar agents after admission. Imaging data included head computerized tomography (CT) scan or magnetic resonance imaging within 24h after onset. Hematoma volume was estimated by the 3D Slicer software package (version 4.10.2; National Institutes of Health, Bethesda, MD, USA).

Well-trained neurologist used the Modified Rankin Scale (mRS) to evaluate patient outcomes through follow-up on telephone at 90 days after onset. The primary outcome was poor outcome at 90 days, defined as mRS>2. This study was approved by the Ethics Committee of the First Affiliated Hospital of Zhengzhou University(Ethics number,2022-KY-1226-002).

### 2.3. Statistical Analysis

Statistical analysis was performed with Statistical Package for Social Sciences (SPSS) version 25.0. Categorical variables were represented by constituent ratios. Non-normally distributed quantitative variables were summarized in form of medians (lower-upper quartiles), and normally distributed quantitative variables were represented as means ± standard deviations. Chi-square test was used to compare categorical variables between two groups. The comparison of continuous variables used Wilcoxon-Mann-Whitney test or independent sample t test. Multivariate logistic regression analysis was performed for significant variables. Receiver operating curve (ROC) was used to determine the best cutoff of 7dBUN/Cr ratio for predicting poor outcome by using the maximum Youden index, and sensitivity and specificity were calculated. A two-tailed *p*<0.05 was identified as statistically significant.

## 3. RESULTS

### 3.1. Baseline Characteristics and Risk Factors of 90-day Poor Outcome After ICH

In this study, a total of 1782 ICH patients were evaluated for eligibility. Finally,211 patients were assessed according to inclusion and exclusion criteria. The flowchart for patient selection is described in **Figure 1**. The demographic, clinical, imaging and laboratory datas are shown in **Table 1**. There were 96 patients (45.5%)with poor outcome, 115 patients (54.5%)with good outcome, and 149 males(70.6%) and 62 females(29.4%), with a mean age of 56 (SD 12) years old. The median hematoma volume was 15.6 (IQR 9.0-21.7) mL. Patients with a poor outcome had a higher median hematoma volume (20.4[15.3,30.0])versus(11.0[6.4,16.8],*p*<0.001), a higher median NIHSS score(11.0[10,13] versus 6.0[4,9], *p*<0.001) and a higher mean ratio of 7dBUN/Cr (19.3[SD 6.1] versus 25.6[SD7.8], *p*<0.001) than those with a good outcome. In addition, univariate analysis also showed that the poor outcome and good outcome groups differed significantly in hypertension history(*p*=0.032), hyperosmolar agents history(*p*=0.001), GCS score(*p*<0.001) and granulocytic lymphocyte ratio(*p*=0.002).

**Figure 1.**
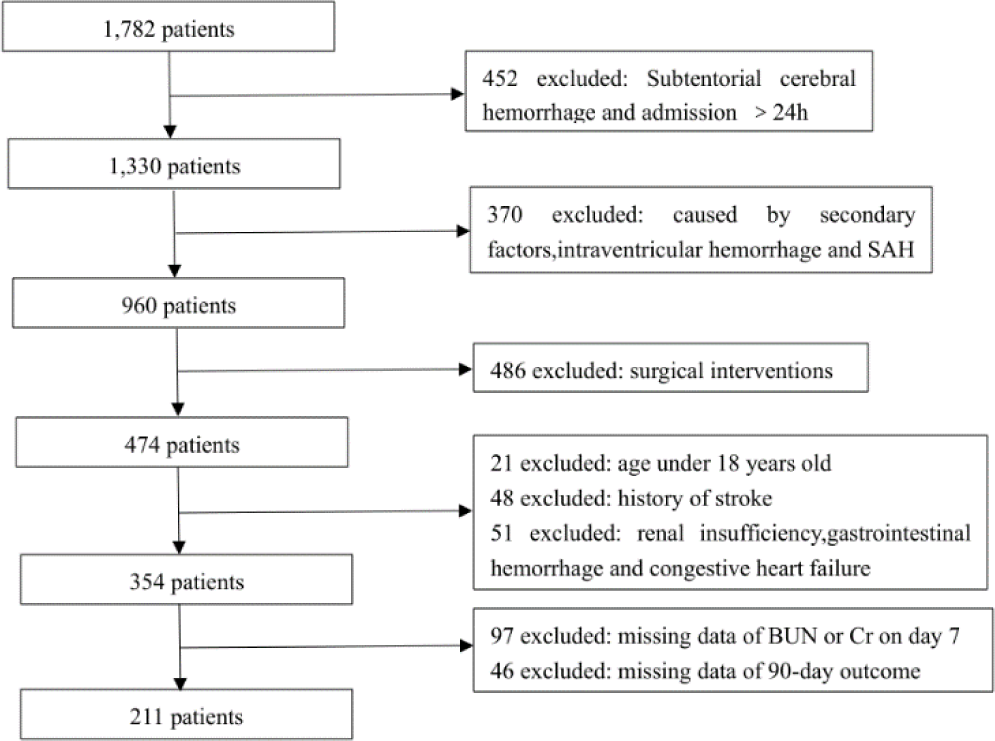
Flowchart for the selection of the study population. Abbreviations: SAH, subarachnoid hemorrhage; BUN, blood urea nitrogen; Cr, creatinine.

**Table 1.** Univariate Analyses of Characteristics of 211 Patients With ICH(n=211)

| Parameter | mRS score0-2<br>( n=115 ) | mRS score 3-6<br>(n=96) | *P* value |
| --- | --- | --- | --- |
| Age(y) | 54.9±11.4 | 57.2±12.7 | 0.162 |
| Male sex | 81(70.4) | 68(70.8) | 0.950 |
| Hypertension | 73(63.5) | 74(77.1) | 0.032* |
| Diabetes mellitus | 16(13.9) | 8(8.3) | 0.204 |
| History of warfarin use | 3(2.6) | 1(1) | 0.746 |
| History of antiplatelet drugs | 10(8.8) | 14(14.7) | 0.180 |
| History of smoking | 35(30.4) | 25(26%) | 0.481 |
| History of drinking | 30(26.1) | 31(32.6) | 0.322 |
| History of hyperosmolar agents | 82(71.3) | 86(89.6) | 0.001* |
| GCS score | 15(14,15) | 14(13,15) | < 0.001* |
| NIHSS score | 6.0(4,9) | 11.0(10,13) | < 0.001* |
| Hematoma volumes(ml) | 11.0(6.4,16.8) | 20.4(15.3,30.0) | < 0.001* |
| Site of ICH |  |  | 0.713 |
| Basal ganglia | 98(85.2) | 87(90.6) |  |
| Thalamic | 8(7) | 4(4.2) |  |
| Lobar | 9(7.8) | 5(5.2) |  |
| Hct | 0.42±0.04 | 0.41±0.04 | 0.445 |
| Platelet(10<sup>9</sup>/L) | 216.1±54.9 | 203.0±68.1 | 0.132 |
| Granulocytic lymphocyte ratio | 5.8(3.4,8.0) | 7.0(5.0,10.0) | 0.002* |
| Low-density lipoprotein | 2.6±0.8 | 2.4±0.8 | 0.102 |
| High-density lipoprotein | 1.2±0.3 | 1.3±0.4 | 0.936 |
| Total cholesterol(mmol/L) | 4.3±0.9 | 4.2±0.9 | 0.134 |
| Albumin(g/L) | 41.5±3.7 | 41±3.6 | 0.500 |
| BUN/Cr ratio on admission | 21.5±7.1 | 20.5±7.0 | 0.228 |
| 7dBUN/Cr ratio | 19.3±6.1 | 25.6±7.8 | < 0.001* |
| Blood sodium(mmol/L) | 139.4±3.5 | 138.7±3.4 | 0.248 |

**Abbreviations:** GCS:Glasgow coma scale,NIHSS:National Institutes of Health Stroke Scale; 7dBUN/Cr ratio: the blood urea nitrogen to creatinine ratio on day 7 after onset, both measured in mg/dL,\* indicates *P* value <0.05.

We generated multivariate logistic regression analysis to identify the independent risk factors that affect the 90-day outcome after ICH. Results are shown in **Table 2**. 7dBUN/Cr ratio was positively correlated with 90-day mRS(OR 1.03,95%CI 1.01-1.05,*p*=0.001), as was NIHSS score(OR 2.09,95%CI 1.60-2.74,*p* < 0.001) and hematoma volume(OR 1.07,95%CI 1.02-1.12,*p*=0.007).

**Table 2.** Multivariate Analysis of Significant Independent Predictors of 90-day Poor Outcome After ICH.

| Patient characteristic | Odds Ratio(95%CI) | *P* |
| --- | --- | --- |
| Hypertension | 1.62(0.63-4.15) | 0.313 |
| History of hyperosmolar agents | 0.59(0.16-2.17) | 0.427 |
| GCS score | 1.10(0.78-1.56) | 0.598 |
| NIHSS score | 2.09(1.60-2.74) | < 0.001* |
| hematoma volumes(ml) | 1.07(1.02-1.12) | 0.007* |
| 7dBUN/Cr ratio | 1.03(1.01-1.05) | 0.001* |
| Granulocytic lymphocyte ratio | 0.96(0.87-1.06) | 0.403 |

Odds ratio is expressed per point of the NIHSS and GCS score, and per 1 ml of ICH volume. \* indicates *P* value <0.05.

### 3.2. Prognostic Predictive Value of 7dBUN/Cr ratio, Hematoma volumes and NIHSS score

ROC curve for 7dBun/Cr ratio, hematoma volume and NIHSS score was used to predict 90-day poor outcome after ICH(**Figure 2**). The 7dBUN/Cr ratio yielded an AUC of 0.753(95%CI,0.685-0.817]). Hematoma volume yielded an AUC of 0.784(95%CI,0.718-0.844), and NIHSS score yielded an AUC of 0.892(95%CI,0.847-0.934). The combined prognostic predictive ability of 7dBUN/Cr ratio, hematoma volume and NIHSS score is improved(AUC 0.922,95%CI 0.887-0.957). The capability of 7dBUN/Cr ratio for predicting poor outcome was equivalent to hematoma volume(*p*=0.507), but not NIHSS score(p<0.001).

**Figure 2.**
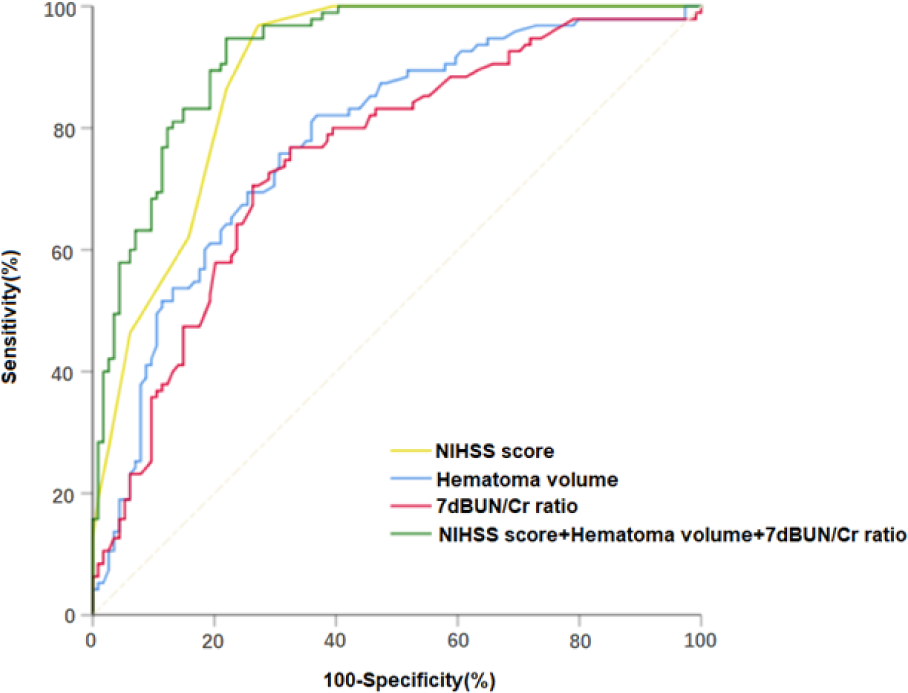
ROC curve for 7dBUN/Cr ratio, hematoma volume and NIHSS score to identify patients with 90-day poor outcome after ICH.

### 3.3 Prognostic Predictive Value of 7dBUN/Cr ratio in patients with hyperosmolar agents after admission

In **Figure 3**, patients with hyperosmolar agents use after admission had a higher level of 7dBUN/Cr ratio(*p*=0.002).

**Figure 3.**
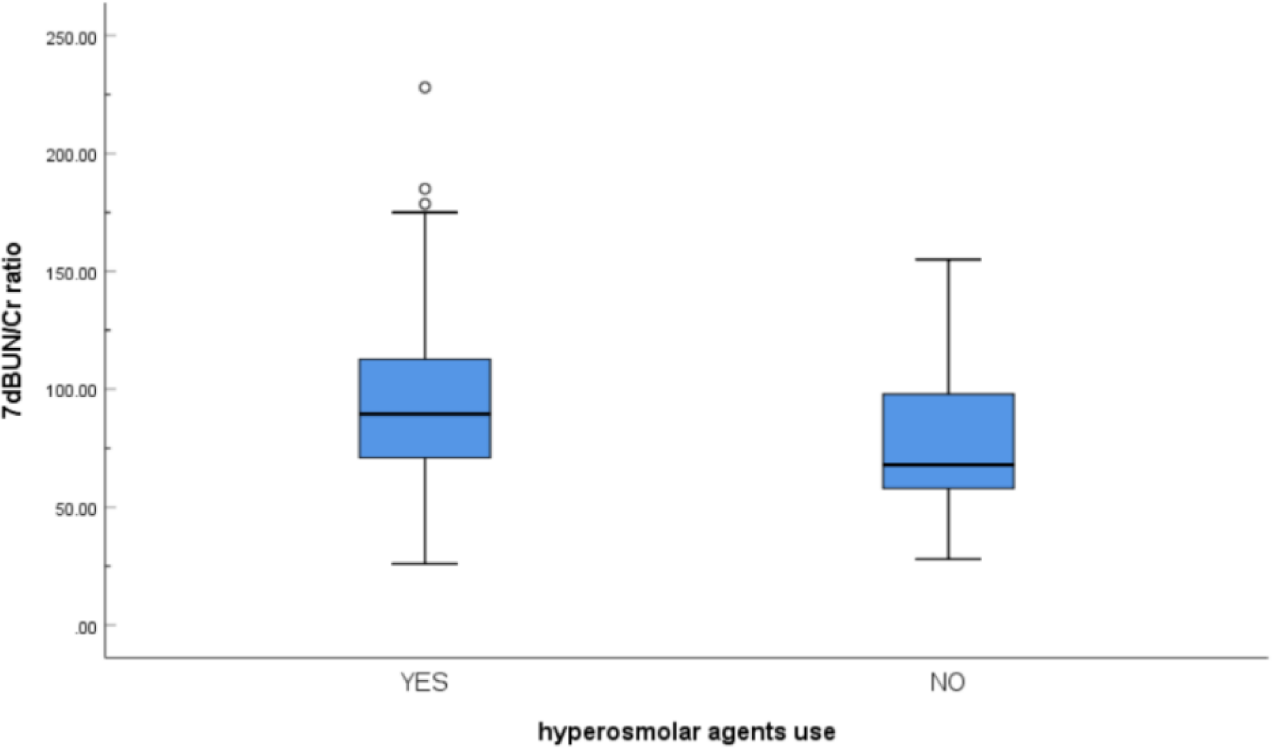
Graph showing difference of 7dBUN/Cr ratio in ICH patients with and without hyperosmolar agents use.7dBUN/Cr ratio are presented as the medians.

In the subgroup of patients treated with hyperosmolar agents, we found that 7dBUN/Cr ratio, NIHSS score, and hematoma volume were also independent risk factors of 90-day poor outcome. ROC curve showed (**Figure 4**)7dBUN/Cr ratio yielded an AUC of 0.751(95%CI,0.676-0.825) to predict 90-day poor outcome.

**Figure 4.**
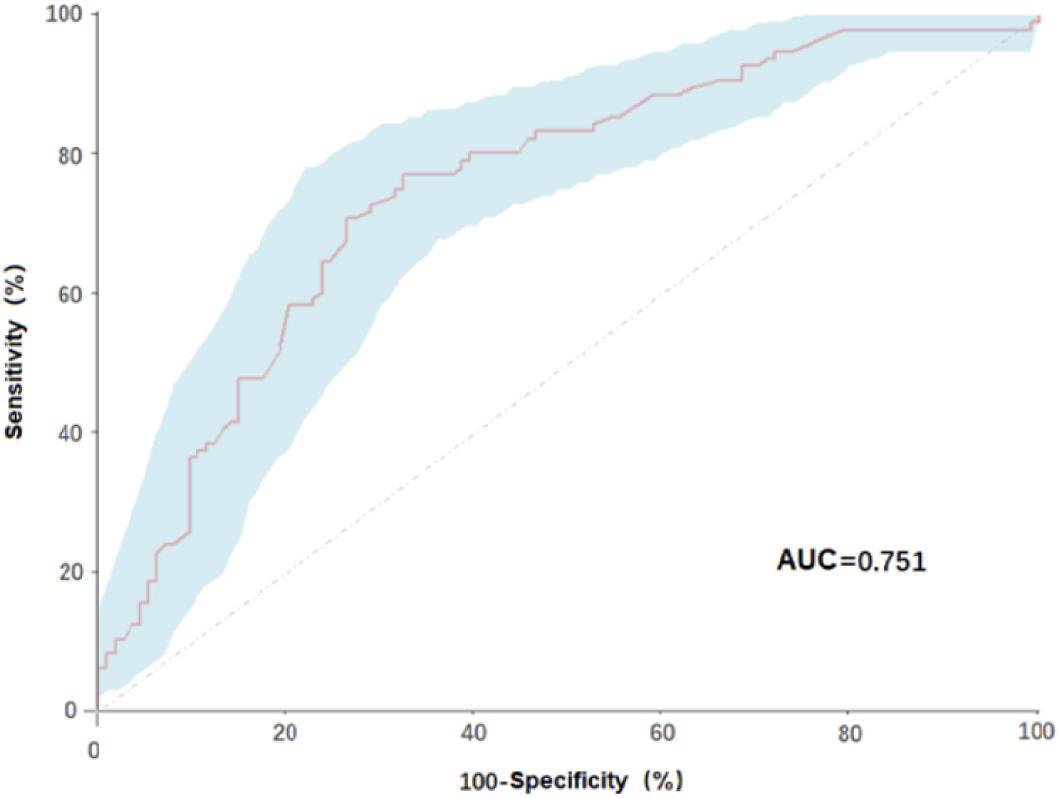
ROC curve with respect to 7dBUN/Cr ratio for predicting 90-day poor outcome of ICH patients with hyperosmolar agents use. AUC means area under curve.

According to the maximum Youden index, the best cutoff point of 7dBUN/Cr ratio was 22 (sensitivity 71.8%, specificity 71.6%).

## 4. DISCUSSION

The evaluation of functional prognosis after ICH is a great challenge. In patients with spontaneous ICH, baseline measures of hemorrahage severity is recommended as an important part in overall evaluation of clinical severity^[2]^ and predicting prognosis. Most baseline severity scores incorporate patient (eg, age), ICH (eg, anatomic location, volume), and clinical deficit characteristics (eg, GCS score, NIHSS score)^[18][19][20]^. However, in addition to baseline severity, some conditions such as treatment-related factors after hospitalization also affect the prognosis of ICH patients.

Dehydration is common in hospitalized stroke patients and is associated with poor outcomes at hospital discharge^[6]^. Blood urea nitrogen-to-creatinine ratio (BUN/Cr) is a biomarker of hydration status. In previous studies, there are three different thresholds to define “dehydration”(BUN/Cr ratios of > 15, > 20, > 25). Elevated BUN/Cr ratio at admission has been confirmed to be associated with poor outcomes in ischemic stroke patients and SAH patients^[7][12][21][22][23]^, but not in ICH patients^[12]^. Our study enrolled 211 patients with spontaneous ICH, and we found BUN/Cr ratio at day 7(7dBUN/Cr) after onset, rather than at admission, was an independent risk factor of 90-day poor outcome, along with NIHSS score and hematoma volume. Well-recognized prognostic predictors for ICH are baseline hematoma volume and status of neurologic function at admission^[25]^. In the study, we found the capability for discriminating risk of poor outcome of NIHSS score(AUC=0.891)was significantly higher than 7dBUN/Cr ratio (AUC=0.751) and hematoma volume (AUC=0.781). While as compared to hematoma volume,7dBUN/Cr ratio displayed similar prognostic predictive abilities in terms of AUC. The result suggests that dehydration during hospitalization is associated with 90-day poor outcome in ICH patients and provides a new indicator for prognosis evaluation after ICH in the future.

In previous study, GCS score, age and hematoma volume were independent prognostic factors in supratentorial ICH patients^[18]^. In our study, both GCS score and NIHSS score were significant factors associated with poor outcome in the univariate analysis. However, multivariate analysis showed that NIHSS score, but not GCS score, was an independent predictor for 90-day poor outcome after ICH. The range of NIHSS score is wider than GCS score, and NIHSS score measures not only the level of consciousness but also the neurological deficits^[26]^. NIHSS score may be a better predictor for outcome than GCS score in both ischemic stroke and ICH. We did not find a correlation between age and prognosis, considering the small number of patients we enrolled and the exclusion of patients with surgery, ventricular hemorrhage and subarachnoid hemorrhage.

In our current results, dehydration is associated with a poor outcome. However, dehydration at admission has been reported to reduce the risk of in-hospital mortality after ICH^[25]^, possibly due to the decrease of blood pressure, hypovolemia and hypernatremia by dehydration, which may increase intravascular osmolality, reduce perihematoma edema and intracranial pressure, but not related to functional outcome. Studies on the relationship between dehydration and ischemic stroke prognosis suggest that dehydration leads to poor prognosis by reducing cerebral perfusion, especially the maintenance of cerebral perfusion during critical periods of brain recovery after stroke^[17]^. Perihematoma hypoperfusion after ICH has been frequently reported, especially delayed perihematomas hypoperfusion after 1 week from onset^[28][29][30]^, and it has been confirmed to be associated with poor outcome in ICH^[31][32]^. We hypothesize that dehydration at admission in ICH patients reduces the risk of mortality and has no relationship with functional outcome, but dehydration after admission may aggravate perihematoma hypoperfusion and then leads to poor long-term functional outcome. This hypothesis needs to be confirmed by prospective studies.

After ICH, acute hematoma and secondary cerebral edema may lead to a rapid increase in intracranial pressure. Hyperosmolar agents, mainly mannitol and hypertonic saline, are the first-line medical therapies for patients with symptomatic brain edema and elevated intracranial pressure^[33]^. In the subgroup comparison, we found that the 7dBUN/Cr ratio was significantly higher in patients with hyperosmolar agents use than those without hyperosmolar agents use, which indicate that dehydration is associated with the application of hyperosmolar agents in ICH patients^[24][34]^. In the hyperosmolar agents subgroup, we found 7dBUN/Cr ratio yielded an AUC of 0.751 to predict 90-day poor outcome and the best cutoff for 7dBUN/Cr ratio was 22 with sensitivity 71.8% and specificity 71.6%. Therefore, 7dBUN/Cr ratio ≥22 appears to be an appropriate threshold for dehydration in ICH patients with hyperosmolar agents use. Current studies have shown that BUN/Cr ratio-based hydration therapy may prevent ischemic stroke progression and improve prognosis^[35][36][37]^. Blood urea nitrogen and creatinine are mainly used to assess renal function and are often used as part of routine examinations in stroke patients. The calculation of the ratio is very simple. Monitoring BUN/Cr ratio may assist in guiding the frequency and time of hypeosmolar agents application and related treatment such as rehydration in clinical practice.

Despite the value of our findings, there are some limitations in our study. First of all, we only enrolled patients with supratentorial cerebral hemorrhage, and excluded patients with the factors which may effect BUN/Cr ratio such as severe infection^[38]^, heart failure^[39]^ and renal insufficiency. These conditions may limit the generalizability of our results to patient populations. Second, our study was retrospective and did not collect datas on fluid therapy after admission. Further prospective studies will be required to confirm these results, and to elucidate the mechanism of dehydration on prognosis of ICH.

## 5. CONCLUSION

Elevated BUN/Cr ratio at day 7 is associated with 90-day poor outcome in ICH patients. Further prospective study will be required to confirm this result and explore the value of BUN/Cr ratio in the application of hyperosmolar agents and hydration therapy.

## Data Availability

All data referred to in the manuscript is available.

